# Preparations of Dutch emergency departments for the COVID-19 pandemic: a questionnaire-based study

**DOI:** 10.1101/2021.04.10.21254878

**Authors:** Rory D O’Connor, Dennis G Barten, Gideon HP Latten

## Abstract

**Background:** The onset of the COVID-19 pandemic was characterized by rapidly increasing patient volumes, which necessitated a swift emergency department (ED) overhaul. Challenges mainly concerned surge capacity, frontline staff protection and the segregation of patients with suspected COVID-19. To date, only few studies have assessed nation-wide ED preparedness for the COVID-19 pandemic. This study aimed to form an overview of preparations that were taken in Dutch EDs during the initial phase of this public health crisis.

**Methods:** This study was designed as a nation-wide, cross-sectional, questionnaire-based study among Dutch hospital organizations with ≥1 ED. The questionnaire was conducted between the first and the second wave of the COVID-19 pandemic in the Netherlands and contained close-ended and open-ended questions on changes in ED infrastructure, ED workforce adaptions and the role of emergency physicians (EPs) in the hospital’s crisis organization.

**Results:** Overall response rate was 79.5%. All EDs had made preparations in anticipation of a possible COVID-19 surge. Treatment capacity was expanded in 69.7% of EDs, with a median increase of 49% (IQR 32.5–72.7%). COVID-19 suspected patients were segregated from non-COVID-19 patients in 86.4% of EDs. Non-COVID-19 patients were more often assessed at alternative locations than patients with suspected COVID-19 infection. In 81.8% of EDs the workforce was expanded, which mainly concerned expansion of nursing staff. A formal role of EPs in the hospital’s crisis organization was reported by 93.9% of EP staffed hospital organizations.

**Conclusion:** All Dutch EDs made preparations for COVID-19 in a short time span and with many uncertainties. Preparations predominantly concerned expansion of treatment capacity and segregation of COVID-19 ED care. EPs had a prominent role, both in direct patient COVID-19 ED care and in the hospitals’ crisis organizations. Although it is vital for EDs to be able to dynamically adapt to community needs, variability of pandemic ED preparedness was high.

## Introduction

Coronavirus disease 2019 (COVID-19) first emerged in Southeast China in December 2019 and was declared a pandemic by the World Health Organization on March 11, 2020.^1^ As COVID-19 spread rapidly around the globe, emergency departments (EDs) within hospitals braced for impact. In the Netherlands, the first case of COVID-19 was identified on February 27, 2020.^2^ As of February 27, 2021, there have been 1,084,021 confirmed cases of infection (of which 24,165 were hospitalized) and 15,543 confirmed COVID-19-deaths.^3^

Emergency medical services and the EDs within hospitals are viewed as the community-based resources responsible for the initial medical response towards any type of disaster, both in the short and long term.^4^ In contrast with sudden-onset events, large-scale infectious outbreaks typically require a prolonged, sustainable response.^4,5^ Since its commencement, the current COVID-19 pandemic was characterized by rapidly increasing patient volumes, which necessitated a swift overhaul of several aspects of ED preparations in Dutch hospitals.^6,7^ Challenges mainly concerned surge capacity, frontline staff protection and the segregation of patients with suspected COVID-19.^8-11^

To date, only few studies have assessed nation-wide ED preparedness for the COVID-19 pandemic. A French questionnaire-based study, conducted during an early stage of the pandemic (March 7 to March 11, 2020), revealed that EDs were poorly prepared.^12^

A similar study from India, limited to academic EDs, showed that 90% of hospitals had developed specific COVID-19 triage systems and that almost 80% established dedicated areas for COVID-19 suspected patients. However, it also revealed that the level of preparedness amongst EDs was highly variable. The authors stated that an individualized strategy for ED preparedness that considers baseline needs and available resources is superior to a blanket strategy for all EDs.^13^

Whilst clinical and intensive care unit (ICU) capacity for COVID-19 in Dutch hospitals were closely monitored and controlled through a national body, there was no guidance on the surge capacity management of EDs.^6^ Consequently, hospitals largely restructured the organization of their EDs on a solitary basis. This study aimed to form an overview of preparations that were taken in Dutch EDs during the initial phase of the COVID-19 pandemic. In addition, it aimed to explore the role of Dutch emergency physicians (EPs) in the hospitals’ crisis organizations.

## Methods

### Setting

The Netherlands is provided with a modern healthcare system with effective primary care and a finely meshed network of specialized acute and critical care facilities, including 83 EDs (Fig 1). The EDs are located within 71 hospital organizations (11 hospital organizations have multiple ED locations). These EDs serve a population of 17.4 million people and have a mean annual attendance rate of 22,500 patients, of which on average 17.4% are self-referred.^14^

**Fig 1.**
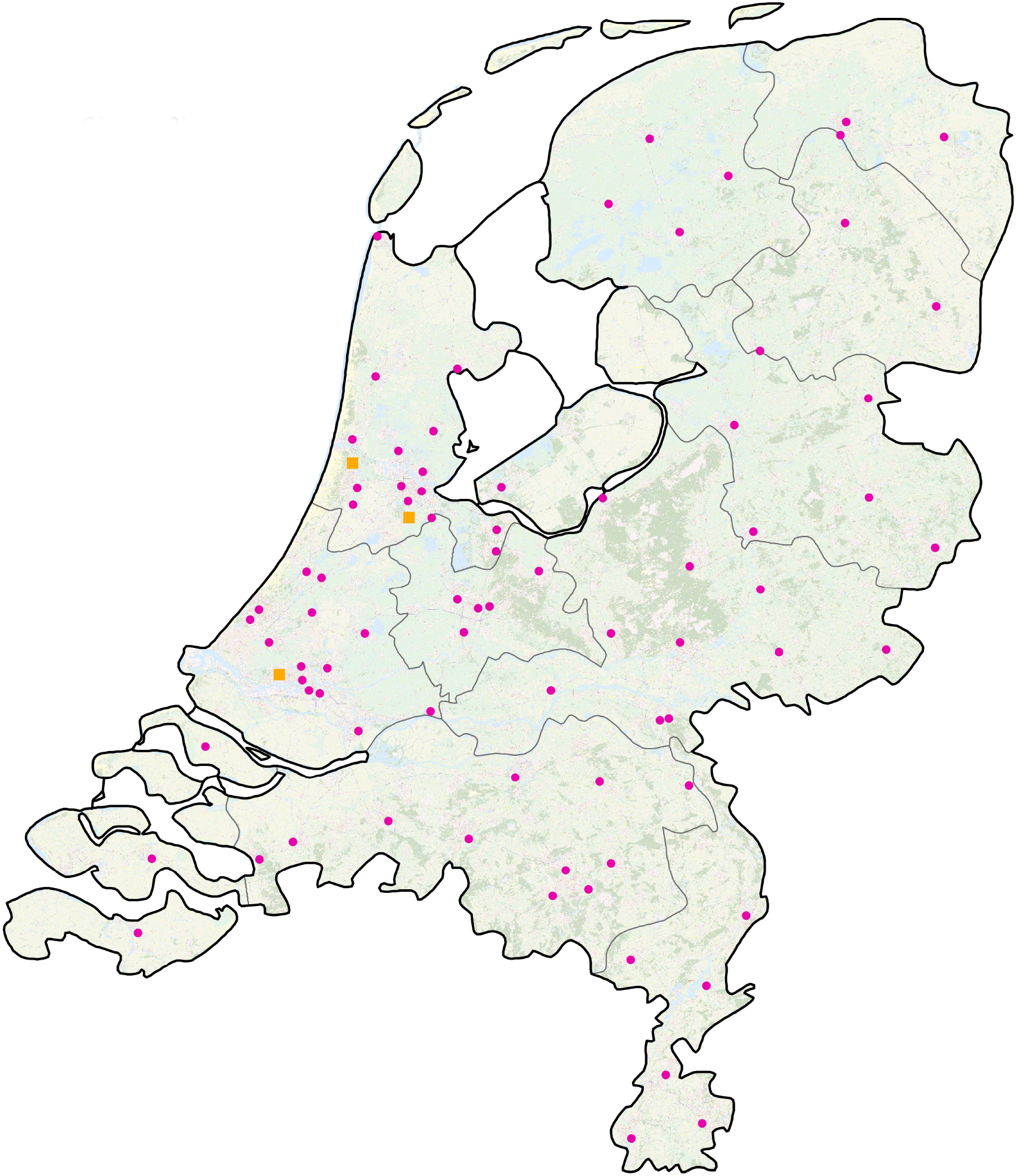
Emergency departments in the Netherlands (June 2020)^15^. - Pink circle: Opened 24 hours, 7 days a week - Yellow square: Opened day and evening, 7 days a week

### Study design

This was designed as a nation-wide, cross-sectional, questionnaire-based study among Dutch hospital organizations with ≥1 ED. For each hospital organization one respondent, consisting of either an EP or an ED manager, received an invitation by email on July 29, 2020. If a respondent did not complete the questionnaire, a reminder was sent every fortnight. The questionnaire could be completed until September 30, 2020. English and Dutch versions of the questionnaire are provided as supplemental file 1 and 2.

Respondents were requested to complete the questionnaire on behalf of the hospital organization where the respondent practiced. When the hospital organization contained multiple EDs, the questionnaire adapted to facilitate information on all EDs. The questionnaire contained 14 close-ended multiple-choice questions and 2 open-ended questions on general ED details and on preparations for a possible surge in COVID-19 patients. Broadly, these questions covered 3 topics: changes in ED infrastructure, ED workforce adaptions and the role of EPs in the hospital’s crisis organization.

### Statistical analysis

All analyses were performed with SPSS version 26 (SPSS Inc., Chicago, USA). Continuous data were reported as means with standard deviation (SD) or as medians with interquartile ranges (IQR). Categorical data were reported as absolute numbers and as valid percentages (to correct for missing data).

All data were collected anonymously. The Strengthening the Reporting of Observational Studies in Epidemiology guidelines was used for reporting this observational study.^16^ The Medical Ethics Committee Zuyderland & Zuyd concluded that the rules of the Medical Research Involving Human Subjects Act (WMO in Dutch) do not apply to this study (METCZ20200130). The study was registered in the Netherlands Trial Register (Trial number NL8818).

## Results

The questionnaire was completed on behalf of 66 (79.5%) out of 83 EDs (Table 1). These EDs served 58 (81.7%) out of 71 hospital organizations, as eight hospital organizations had multiple ED locations.

**Table 1.**
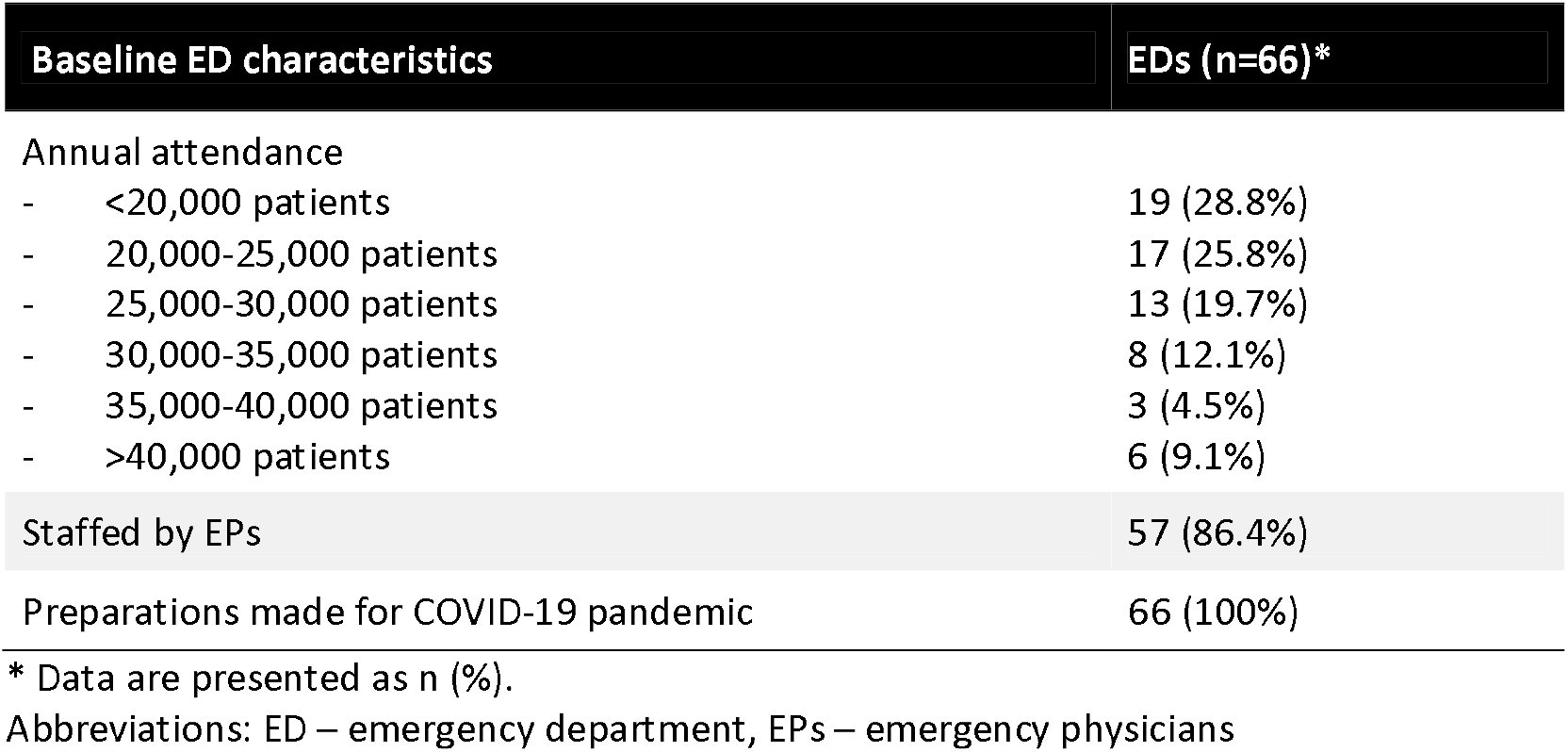

| Table 1 – Baseline ED characteristics | EDs (n=66)* |
| --- | --- |
| Annual attendance |  |
| - <20,000 patients | 19 (28.8%) |
| - 20,000-25,000 patients | 17 (25.8%) |
| - 25,000-30,000 patients | 13 (19.7%) |
| - 30,000-35,000 patients | 8 (12.1%) |
| - 35,000-40,000 patients | 3 (4.5%) |
| - >40,000 patients | 6 (9.1%) |
| Staffed by EPs | 57 (86.4%) |
| Preparations made for COVID-19 pandemic | 66 (100%) |

\* Data are presented as n (%).
Abbreviations: ED – emergency department, EPs – emergency physicians

Pre-COVID, the majority of the EDs had an annual attendance rate of fewer than 30.000 patients per year and 86.4% of EDs were staffed by EPs.

All participating EDs had made preparations in anticipation of a possible surge of COVID-19 patients. The date when these preparations were finalized varied between February 24 and May 5, 2020

### Changes in ED infrastructure

Before the COVID-19 pandemic, the median number of ED treatment spaces was 17 (IQR 12–21) (Table 2). Treatment capacity was expanded in 46 (69.7%) EDs. The median number of additional treatment spaces was 8 (IQR 4-10), which equals to a median increase of 49% (IQR 32.5–72.7%). Explanations for not increasing the ED capacity included previous reduction of ED utilization by several logistic alterations (15.2%), being designated as a non-COVID-19 ED (6.1%), and the inability to expand ED treatment spaces due to isolation measures demanding more space per patient (4.5%). Logistic alterations to usual practice included the redirection of low-acuity ED visits, such as minor traumatic injuries, to outpatient departments in 41 (62.1%) EDs. Furthermore, 12 (18.1%) EDs effectuated a faster admission process to hospital wards and intensive care units, therewith shortening ED length of stay.

**Table 2.**

| Table 2 – Changes in ED infrastructure | EDs* |
| --- | --- |
| Pre-pandemic treatment spaces | 17 (12-21) |
| Treatment spaces increased during pandemic | 46 (70.0%) |
| – Additional treatment spaces | 8 (4-10) |
| No increase in treatment spaces | 20 (30.3%) |
| – Logistic alterations to usual ED practice | 10 (15.2%) |
| – non-COVID-19 hospital | 4 (6.1%) |
| – Expansion not feasible | 3 (4.5%) |
| – Other | 3 (4.5%) |
| Segregation of COVID-19 ED care | 57 (86.4) |
| Location of COVID-19 ED care |  |
| – Original ED only | 39 (68.4%) |
| – Original ED and other location | 14 (24.6%) |
| – Other location only | 4 (7.0%) |
| Location of non-COVID-19 ED care |  |
| – Original ED only | 27 (47.4%) |
| – Original ED and other location | 23 (40.4%) |
| – Other location only | 7 (12.3%) |
| Screening for COVID-19 before ED entry performed with |  |
| – Symptom-based screening list only | 43 (75.4%) |
| – Symptom-based screening list and radiological imaging (Chest X-ray or CT) | 13 (22.8%) |
| – Chest CT only | 1 (1.8%) |

\* Data are presented as median (IQR), or n (%).
Abbreviations: ED – emergency department

COVID-19 suspected patients were segregated from non-COVID-19 patients in 57 (86.4%) EDs. In the majority (68.4%) of EDs, this was organized within the original ED space. The alternative locations used by the remaining EDs can be found in supplemental file 3. In most (75.4%) EDs, the suspicion of COVID-19 was established using a symptom-based checklist only.

In 53 (80.3%) hospital organizations, one or more of the implemented measures for the pandemic were intended to be maintained (Supplemental file 4). These included improved infection prevention in 13 (22.4%), improved interdisciplinary collaboration in 13 (22.4%), permanent adjustments to segregate possibly contagious patient categories in 10 (17.2%) and permanent redirection of low-urgent patient categories 8 (13.8%) hospital organizations.

### ED workforce adaptations

In 54 (81.8%) EDs the workforce was expanded (Table 3). In all of these EDs nursing staff was expanded by deploying both additional specialized ED nurses (53.0%) as well as nurses from other departments (60.6%). A large variety of physicians were directly involved in COVID-19 ED care, of which emergency medicine (86.4%), internal medicine (84.8%) and pulmonology (81.8%) were involved most frequently. In 21 (31.8%) EDs, the additional workforce consisted of nurses and physicians only, whereas other disciplines were also deployed in the remaining 45 (68.2%) EDs.

**Table 3.**

| Table 3 – ED workforce adaptations | EDs* |
| --- | --- |
| Expansion of nursing staff | 54 (81.8%) |
| - Additional ED nurses | 35 (53.0%) |
| - Additional non-ED nurses | 40 (60.6%) |
| - Specialties involved in ED COVID-19 care |  |
| Emergency medicine | 57 (86.4%) |
| - Internal medicine | 56 (84.8%) |
| - Pulmonology | 54 (81.8%) |
| - Anesthesiology | 26 (39.4%) |
| - Geriatrics | 24 (36.4%) |
| - Surgery | 23 (34.8%) |
| - Neurology | 22 (33.3%) |
| - Cardiology | 20 (30.3%) |
| - Pediatrics | 20 (30.3%) |
| - Otolaryngology/ENT | 18 (27.3%) |
| - Gastro-enterology | 16 (24.2%) |
| - Orthopedics | 14 (21.2%) |
| - Urology | 12 (18.2%) |
| - Dermatology | 6 (9.1%) |
| - Primary care | 6 (9.1%) |
| - Plastic surgery | 6 (9.1%) |
| - Rheumatology | 5 (7.6%) |
| - Gynecology | 4 (6.1%) |
| - Other | 16 (24.2%) |
| - Other disciplines | 44 (66.7%) |
| - Physician assistants | 17 (25.8%) |
| - Medical interns | 17 (25.8%) |
| - Doctor's assistants | 15 (22.7%) |
| - Surgery assistants | 14 (21.2%) |
| - Anesthetic nurses | 9 (13.6%) |
| - Plaster technicians | 9 (13.6%) |
| - Other** | 7 (10.6%) |

\* Data are presented as n (%). \*\* Volunteers, medical students
Abbreviations: ED – emergency department, ENT – ear nose throat

### Role of EPs in the crisis organization

EPs were staffed in 49 (84.5%) hospital organizations. In all of these hospital organizations, EPs were directly involved in the assessment and treatment of COVID-19 patients. In addition, in hospital organizations staffed by EPs, EPs had a coordinating role in the ED in 44 (89.8%) and were involved in triage or segregation of COVID-19 suspected patients in 40 (81.6%) hospital organizations. A formal role of EPs in the hospital’s crisis organization was reported in 46 (93.9%) hospital organizations. An EP was member of the strategic crisis team in 19 (38.8%) hospital organizations and of the operational crisis team in 34 (69.4%) hospital organizations.

### Crowding

The majority (52%) of hospital organizations did not experience crowding during the first COVID-19 surge. Occasional crowding was reported by 24 (41%) and no crowding by 30 (52%) hospital organizations.

## Discussion

This questionnaire-based study aimed to provide an overview of preparations of Dutch EDs for the initial phase of the COVID-19 pandemic. With a response rate of 79.5% of EDs, the results are representative for all Dutch EDs.

All participating EDs made preparations for a surge in COVID-19 patients. Treatment capacity was expanded in almost 70% of the participating EDs, with a median increase in treatment spaces of 50%. COVID-19 patients were segregated from non-COVID-19 patients in 86.4% of EDs, and ED workforce was expanded in 81.8% of EDs. EPs were directly involved in the care for COVID-19 patients in all EDs and had a prominent role in the crisis organization in 93.9%.

The COVID-19 pandemic forced EDs to make drastic organizational changes in a very short time span. At the time, it was unclear for EDs if they would be either sufficient or even necessary.^8^ In the Netherlands, there was national guidance on clinical and ICU capacity.^6^ Remarkably, there was no alignment or general advice on the expansion of ED capacity. This is reflected by the heterogeneity of the results of this study. Although more alignment between EDs may be desirable, the solitary surge capacity plans worked for most EDs as the majority reported no or occasional crowding. In this perspective, it is important to acknowledge that ED surge capacity planning should be accommodated to specific hospital characteristics and leave room for improvisation, even when there is national guidance. During the first wave of the COVID-19 pandemic, regions within the Netherlands differed considerably with regards to COVID-19 infection rates. This may have influenced the burden on EDs and could in part explain the heterogeneity between pandemic ED approaches as well.

The COVID-19 pandemic may have changed ED care forever, as some adaptions will remain. E-health applications have flourished and there is more focus on getting the right care in the right place.^17^ Some patient categories do not necessarily need ED care, but may receive safe and efficient care at another location. Furthermore, this public health crisis has shown the importance of a strong emergency and critical care system, where a certain degree of overcapacity may be pivotal for an effective response. As this pandemic is ongoing, surge capacity models that allow flexibility to some extent may be most useful.^7,9,18^ Hospital capacity is dynamic and highly dependent on the occupancy of available resources.^19^ At times when the pressure on ED care is temporarily lower, capacity could be used for non-urgent care and vice versa. This way, hospitals could timely anticipate to community demands.

Close collaboration within hospitals has always been of vital importance. As shown by our results, virtually all medical disciplines were deployed in the EDs during the pandemic. Although this survey did not examine the quality of inter-disciplinary collaboration, multiple respondents greatly valued the unique situation where all kinds of disciplines worked closely together. It may not come as a surprise that EPs, internists and pulmonologists were involved in COVID-19 ED care. However, EPs also played an important role in ED coordination and triage. Furthermore, EPs took vital positions in the hospitals’ crisis organizations, underling the necessity of experienced staff members working specifically in the ED.

The present study is not without limitations. First, this was a retrospective questionnaire-based study filled in by one respondent per ED, who may also have been the most involved professionals in crisis management in these EDs. Second, hospitals all around the world experienced reduced utilization of emergency services during the pandemic.^20^ This phenomenon, which is not yet completely understood, may have salvaged EDs which could have suffered from overcrowding without the reduction of non-COVID-19 ED care. In other words, the pandemic approaches of these EDs may not be as successful in other crisis situations. Finally, the results of this study may not apply to EDs in other healthcare systems, specifically systems without a strong primary care system functioning as gatekeepers for the hospitals.

## Conclusion

This study showed that all Dutch EDs made preparations for COVID-19 in a short time span and with many uncertainties. Preparations primarily included the expansion of treatment capacity and the segregation of COVID-19 care. EPs had a prominent role, both in direct patient COVID-19 ED care and in the crisis organizations of hospitals. Although it is vital for EDs to be able to dynamically adapt to community needs, variability of pandemic ED preparedness was high.

## Data Availability

All relevant data are within the manuscript and its Supporting Information files.

## Abbreviations

EP: Emergency physician
ED: Emergency department
ICU: Intensive care unit
IQR: Interquartile Range

## Acknowledgements

We would like to thank all participating EDs for their participation in this study. We also want to thank the Dutch Society of Emergency Physicians for its support in distributing the questionnaire among the EDs.

## Collaborators

L.M. Esteve Cuevas, M.L. Ridderikhof, dr W.A.M.H. Thijssen, R.R. Pigge, N.E. Mullaart-Jansen, R.J.C.G. Verdonschot, V. Brown, G. van Woerden, E.L. Janssens, B.Y.M. van der Kolk, B. de Groot, F. Derkx-Verhagen, W.P. Poortvliet, H. Lameijer, Y. Schoon, J. Holkenborg, L.E. Kerkvliet, M.S.A. de la Fosse, E. ter Avest, MD, PhD., K. Azijli, S. Postma, J.M. van Lieshout, B. Vlaming, C. Kok, M. Maltha, R. Lulf, R.J.L. Boden, A.E. Boendermaker, J.L.P. Kuijten, J.L. van der Meer, K. van den Broek, L. Jansen, M.J. Meijer, T.B. Nanlohij, D.J.R. Keereweer, A.G. Pol, T.J. Oosterveld-Bonsma, J.M. Huttenhuis, G.B. Spijkers

## Conflicts of interest

There are no conflicts of interest.

## Supporting information

**S1 Questionnaire English**

**S2 Questionnaire Dutch**

**S3 Alternative locations of emergency care**

**S4 Measures intended to be maintained**

## Notes

### Competing Interest Statement

The authors have declared no competing interest.

### Clinical Trial

The study was registered in the Netherlands Trial Register (Trial number NL8818)

### Funding Statement

No external funding was received.

### Author Declarations

The Medical Ethics Committee Zuyderland & Zuyd concluded that the rules of the Medical Research Involving Human Subjects Act (WMO in Dutch) do not apply to this study (METCZ20200130).

