## Supplemental file 1 for "Preparations of Dutch emergency departments for the COVID-19 pandemic: a questionnaire-based study"

### Supplemental file 1 - Questionnaire (English)

1. What is the annual ED attendance?
   - <20.000 patients
   - 20.000-25.000 patients
   - 25.000-30.000 patients
   - 30.000-35.000 patients
   - 35.000-40.000 patients
   - >40.000 patients
2. Is the ED staffed by EPs?
   - Yes
   - No
3. How many treatment places does the ED have?
4. Have there been preparations with regards to the ED in anticipation of COVID-19 patients?
   - Yes
   - No
5. On what date were these preparations implemented? .. - ..-2020
6. Was the number of ED treatment places expanded because of the COVID-19 pandemic?
   - Yes, it was expanded to … ED treatment places (including treatment places for delivering standard emergency care and COVID-19 emergency care).
   - No. Could you elaborate why the number of ED treatment places was not expanded: …
7. Were arrangements made to relieve the ED?
   - Yes. Could you elaborate on these arrangements? …
   - No
8. Was the flow of COVID-19 patients needing emergency care segregated from the flow of standard emergency department patients?
   - No
   - Yes
     - Where did the emergency care for COVID-19 patients take place?
       - Original ED only
       - Original ED and other location: Which was …
       - Only other location: Which was …
     - Where did the emergency care for COVID-19 patients take place?
       - Original ED only
       - Original ED and the following other location: Which was …
       - Only other location: Which was …
9. If COVID-19 care was segregated, screening for COVID-19 before ED entry was performed by:
   A. Symptom-based screening list:
      - Yes
      - No
   B. Chest X-ray
      - Yes
      - No
   C. Chest CT
      - Yes
      - No
10. Was ED personnel capacity increased because of the COVID-19 pandemic?
    - No
    - Yes
      A. Additional ED nurses
         - Yes
         - No
      B. Additional non-ED nurses
         - Yes
         - No
11. Were the following specialties involved in the COVID-19 care in the ED?
    A. Anaesthesiology
       - Yes
       - No
    B. Cardiology
       - Yes
       - No
    C. Dermatology
       - Yes
       - No
    D. Emergency medicine
       - Yes
       - No
    E. Gastro-enterology
       - Yes
       - No
    F. Geriatrics
       - Yes
       - No
    G. Gynaecology
       - Yes
       - No
    H. Internal medicine
       - Yes
       - No
    I. Neurology
       - Yes
       - No
    J. Orthopaedics
       - Yes
       - No
    K. Otolaryngology/ENT
       - Yes
       - No
    L. Paediatrics
       - Yes
       - No
    M. Plastic surgery
       - Yes
       - No
    N. Primary care
       - Yes
       - No
    O. Pulmonology
       - Yes
       - No
    P. Rheumatology
       - Yes
       - No
    Q. Surgery
       - Yes
       - No
    R. Urology
       - Yes
       - No
    S. Other
       - Yes, could you specify? …
12. Were the following disciplines involved in the COVID-19 care in the ED?
    A. Anesthetic nurses
       - Yes
       - No
    B. Medical interns
       - Yes
       - No
    C. Doctors’ assistants
       - Yes
       - No
    D. Plaster technicians
       - Yes
       - No
    E. Surgery assistants
       - Yes
       - No
    F. Physician assistants
       - Yes
       - No
    G. Other. Coud you specify? …
13. Did EPs have a role in COVID-19 related emergency care?
    - No
    - Yes
      - EPs were directly involved in the assessment and treatment of COVID-19 patients
        - Yes
        - No
      - EPs had a coordinating role regarding COVID-19 ED care.
        - Yes
        - No
      - EPs were involved in triage or segregation of COVID-19 suspected patients
        - Yes
        - No
14. Did EPs have a formal role in the hospital’s crisis organization?
    - No
    - Yes
      - EPs were member of the strategic crisis team
        - Yes
        - No
      - EPs were member of an operational crisis team
        - Yes
        - No
      - They were involved in triage or segregation of COVID-19 suspected patients
        - Yes
        - No
15. Has overcrowding of the ED occurred during the first surge of COVID-19 patients?
    - Yes, occasionally
    - Yes, multiple times a week
    - Yes, daily
    - Yes, multiple times a day
    - No
16. Do you expect that measures that were implemented in the ED due to the COVID-19 pandemic will be instated permanently?
    - Yes, could you elaborate on these measures? …
    - No
