## Supplemental file 2 for "Preparations of Dutch emergency departments for the COVID-19 pandemic: a questionnaire-based study"

### Supplemental file 2 - Questionnaire (Dutch)

1. Hoeveel patiënten ontvangt uw SEH ongeveer per jaar?
   - <20.000 patiënten
   - 20.000-25.000 patiënten
   - 25.000-30.000 patiënten
   - 30.000-35.000 patiënten
   - 35.000-40.000 patiënten
   - >40.000 patiënten
2. Zijn er SEH-artsen KNMG werkzaam in uw ziekenhuis
   - Ja
   - Nee
3. Hoeveel behandelplekken had uw SEH voorafgaande aan COVID-19?
4. Zijn/waren er op uw SEH veranderingen doorgevoerd vanwege COVID-19?
   - Ja
   - Nee
5. Op welke datum zijn de eerste veranderingen geëffectueerd? ..-..-2020
6. Was het aantal behandelplekken op de SEH uitgebreid in het kader van de COVID-19 pandemie?
   - Ja, dit is uitgebreid naar … behandelplekken (Dit aantal betreft zowel de behandelplekken voor standaard zorg als COVID-19 zorg)
   - Nee, kunt u toelichten waarom het aantal SEH behandelplekken niet is uitgebreid?
7. Zijn/waren er binnen het ziekenhuis afspraken gemaakt om de SEH te ontlasten?
   - Ja, kunt u deze afspraken nader toelichten?
   - Nee.
8. Wordt/werd de acute patiëntenstroom van COVID-19 verdachte patiënten gescheiden van non-COVID patiënten in uw ziekenhuis?
   - Nee
   - Ja
     A. Waar vond de SEH zorg voor COVID-verdachte patiënten plaats?
        - Alléén op de reguliere SEH
        - Op de reguliere SEH én op een andere locatie binnen het ziekenhuis, namelijk …
        - Alléén op een andere locatie binnen het ziekenhuis, namelijk …
     B. Where did the emergency care for COVID-19 patients take place?
        - Alléén op de reguliere SEH
        - Op de reguliere SEH én op (een) andere locatie(s) binnen het ziekenhuis, namelijk …
        - Alléén op een andere locatie binnen het ziekenhuis, namelijk …
9. Op welke manier werd bij de instroom naar de SEH bepaald of een patiënt al dan niet COVID verdacht was?
   - Aan de hand van screeningslijst met klachten
     - Ja
     - Nee
   - X-thorax
     - Ja
     - Nee
   - CT-thorax
     - Ja
     - Nee
10. Was er uitbreiding van de hoeveelheid personeel op de SEH?
    - Nee
    - Ja
      A. Is het aantal SEH verpleegkundigen uitgebreid?
         - Ja
         - Nee
      B. Is het aantal verpleegkundigen op de SEH uitgebreid met verpleegkundigen van andere afdelingen?
         - Ja
         - Nee
11. Waren de volgende specialismen betrokken bij de COVID-19 zorg op de SEH?
    A. Anesthesie
       - Ja
       - Nee
    B. Cardiologie
       - Ja
       - Nee
    C. Dermatologie
       - Ja
       - Nee
    D. Spoedeisende Geneeskunde
       - Ja
       - Nee
    E. Maag-, Darm- en Leverziekten
       - Ja
       - Nee
    F. Geriatrie
       - Ja
       - Nee
    G. Gynaecologie
       - Ja
       - Nee
    H. Interne geneeskunde
       - Ja
       - Nee
    I. Neurologie
       - Ja
       - Nee
    J. Orthopedie
       - Ja
       - Nee
    K. Keel-neus-oorheelkunde
       - Ja
       - Nee
    L. Kindergeneeskunde
       - Ja
       - Nee
    M. Plastische Chirurgie
       - Ja
       - Nee
    N. Huisartsgeneeskunde
       - Ja
       - Nee
    O. Longziekten
       - Ja
       - Nee
    P. Rheumatologie
       - Ja
       - Nee
    Q. Chirurgie
       - Ja
       - Nee
    R. Urologie
       - Ja
       - Nee
    S. Anders
       - Ja, namelijk …
12. Waren de volgende functies op de SEH betrokken bij de directe zorg voor COVID-19 verdachte patiënten?
    A. Anesthesiemedewerkers
       - Ja
       - Nee
    B. Coassistenten
       - Ja
       - Nee
    C. Dokters assistenten
       - Ja
       - Nee
    D. Gipsverbandmeesters
       - Ja
       - Nee
    E. Operatie assistenten
       - Ja
       - Nee
    F. Physician Assistants
       - Ja
       - Nee
    G. Anders, namelijk: …
13. Heeft de SEH-arts KNMG een rol in de COVID-19 gerelateerde zorg op de SEH?
    - Nee
    - Ja, zij hadden zij de volgende rol(len):
      A. Zij waren betrokken bij de directe opvang en behandeling van patiënten.
         - Ja
         - Nee
      B. Zij hadden een rol als medisch coördinator van de COVID-19 SEH
         - Ja
         - Nee
      C. Zij zorgden voor de scheiding/triage van acute patiëntenstroom in COVID-19 verdacht en non-COVID-19 verdacht
         - Ja
         - Nee
14. Heeft de SEH-arts KNMG een formele rol in de COVID crisisorganisatie gehad ?
    - Ja, zij hadden zij de volgende rol(len):
      A. De SEH-arts maakte deel uit van het crisisbeleidsteam
         - Ja
         - Nee
      B. De SEH-arts maakte deel uit van een operationeel team
         - Ja
         - Nee
      C. De SEH-arts gaf input vanaf de werkvloer naar het management
         - Ja
         - Nee
    - Nee
15. Is er in het voorjaar van 2020 tijdens het hoogtepunt van de pandemie in Nederland een moment (of momenten) geweest waarop de capaciteit van de SEH tekortschoot?
    - Ja, een enkele keer
    - Ja, meerdere keren per week
    - Ja, dagelijks
    - Ja, meermaals per dag
    - Nee
16. Zijn er veranderingen op de SEH doorgevoerd in het kader van de COVID-crisis, waarvan u denkt dat deze na de crisis blijven bestaan?
    - Ja, kunt u aangeven welke maatregelen dat zijn?
    - Nee
