## Supplemental file 3 for "Preparations of Dutch emergency departments for the COVID-19 pandemic: a questionnaire-based study"

### Supplemental file 4 – Alternative locations of emergency care

| Location for COVID-19 ED care | EDs^*^ |
| --- | --- |
| Tent or decontamination unit adjacent to the ED | 7 (10.7) |
| COVID outpatient department (in case of stable vitals) | 4 (6.1%) |
| Space in specialised facility (such as minor surgery and endoscopy unit) | 1 (1.5%) |
| Fast lane cardiology clinic | 2 (3.0%) |
| Fast lane respiratory clinic | 2 (3.0%) |
| Other hospital** | 2 (3.0%) |
| COVID screening department | 1 (1.5%) |
| Pulmonologists and intensivists did consultations of patients at General Practice Cooperative adjacent to ED and decided to either admit to ward or send patient home | 2 (3.0%) |

* numbers are presented as n (%)

**Hospital was designated as non-COVID-19 location

| Location for non-COVID ED care | EDs^*^ |
| --- | --- |
| ED Patients with stable vitals at outpatient department** | 38 (57.8%) |
| Other non-COVID ED location was set up within hospital | 7 (10.6%) |
| Assessment in department adjacent to ED | 11 (16.7%) |
| Other hospital*** | 2 (3.03%) |
| Tent | 2 (3.03%) |
| ED care for specific patient groups |  |
| Children in paediatric ward | 10 (15.2%) |
| Thrombolysis eligible stroke patients in stroke unit | 3 (4.5%) |

* numbers are presented as n (%)

****** These patients included children and for example patients suffering minor traumatic injuries, oncologic disease and deep venous thrombosis.

***Hospital was designated as COVID-19 location
