## Supplemental file 4 for "Preparations of Dutch emergency departments for the COVID-19 pandemic: a questionnaire-based study"

### Supplemental file 4 – Measures implemented due to COVID-19 that are intended to be maintained.

| Measure | Hospital organizations* |
| --- | --- |
| Improved infection prevention | 13 (22.4%) |
| Improved interdisciplinary collaboration | 13 (22.4%) |
| Permanent adjustments to segregate possibly contagious patient categories | 10 (17.2%) |
| Permanent redirection of low-urgent patient categories | 8 (13.8%) |
| Expansion of staff | 5 (8.6%) |
| Dynamic ED capacity model with adjacent hospital department (i.e. acute medical unit) | 4 (6.9%) |
| E-health/telemedicine | 4 (6.9%) |
| Permanent expansion of capacity | 4 (6.9%) |
| Improved throughput | 4 (6.9%) |
| More prominent role of emergency department | 3 (5.2%) |
| Extra diagnostic modalities | 3 (5.2%) |
| Improved transmural collaboration | 2 (3.4%) |
| Improved security (of staff and facility) | 1 (1.7%) |
| National coordination of clinical capacity | 1 (1.7%) |

* Data are presented as n (%)

Abbreviations: ED – emergency department
